# Transmission risk of SARS-CoV-2 on airplanes and high-speed trains

**DOI:** 10.1101/2020.12.21.20248383

**Authors:** Maogui Hu, Jinfeng Wang, Hui Lin, Corrine W Ruktanonchai, Chengdong Xu, Bin Meng, Xin Zhang, Alessandra Carioli, Yuqing Feng, Qian Yin, Jessica R Floyd, Nick W Ruktanonchai, Zhongjie Li, Weizhong Yang, Andrew J Tatem, Shengjie Lai

## Abstract

Modern transportation plays a key role in the long-distance and rapid spread of SARS-CoV-2. However, little is known about the transmission risk of SARS-CoV-2 on confined vehicles, such as airplanes and trains. Based on the itinerary and epidemiological data of COVID-19 cases and close contacts among 9,265 airline passengers on 291 airplanes and 29,335 passengers on 830 high-speed trains in China from December 20, 2019 to March 17, 2020, we estimated that the upper bound of overall attack rate of COVID-19 among passengers was 0.60% (95% confidence interval: 0.43%-0.84%) for airplanes and 0.35% (0.28%-0.44%) for trains departing from Wuhan before its lockdown, respectively. The reproduction number during travel ranged from 0.12 to 0.19 on airplanes and from 0.07 to 0.12 on trains, with the risk varying by seat distance from the index case and joint travel time, but the difference in risk was not significant between the types of aircraft and train. Overall, the risk of SARS-CoV-2 transmission on planes and high-speed trains with high efficiency air filtration devices was relatively low. Our findings improve understanding of COVID-19 spread during travel and may inform response efforts, such as lifting travel restrictions, and resuming transportation in the pandemic.

## Main

The current COVID-19 pandemic has affected nearly all countries across the world within a matter of months, causing tens of millions of infections and hundreds of thousands of deaths in 2020 ^1^. Large-scale population movements through long-distance travel are one of the most important reasons for the rapid spread of SARS-CoV-2 across regions and countries ^2,3^. Many studies have shown that SARS-Cov-2 transmission in a city or country was initiated by the virus introduced by domestic or international travellers from epidemic regions, and the number of imported cases by country in the early stage of the pandemic has a significant correlation with the volume of travellers from other epidemic areas ^4-8^. Towards this, many domestic and international travel routes have been restricted or suspended to contain the further spread of SARS-CoV-2 ^9^.

Airplanes and trains have become some of the most important modes of transportation for long-distance travel within and across many countries, particular in Europe, North America, and East Asia. According to the World Bank statistics, the number of global air passengers reached 4.2 billion in 2018 ^10^, and about 8 billion passengers travelled on national railway networks in the countries of the European Union in 2018, with 2 billion passengers travelling by high-speed trains in mainland China ^11,12^. Due to travel restrictions during the pandemic, the transport industry has been hit hard. For countries trying to resume socioeconomic activities and transportation, the spread of SARS-CoV-2 through travellers may lead to another resurgence and lockdown if the risk is not fully recognised and addressed. For example, according to the Flightradar24 (www.flightradar24.com), there were as many as 6972 flights in the skies above North America two days ahead of the Thanksgiving holiday in the United States (US) in 2020, which were more than that in 2018, despite recommendations from the US Centers for Disease Control and Prevention (CDC) to reduce non-essential travelling during the winter wave of COVID-19^13^.

Airlines and railways preparing to resume timetables have been also struggling to implement social distancing guidelines, such as leaving the middle seats unoccupied on airplanes or trains to reduce COVID-19 risk ^14,15^. These measures were in response to studies suggesting increased SARS-CoV-2 transmission among passengers in confined spaces ^16-22^; for example, Speake et al. (2020) found flight-associated SARS-CoV-2 transmission on a domestic flight route in Australia and reported that the secondary infectious rate was about 9.8% in the mid-portion of the cabin ^16^. On the contrary, some research observed that the risk of SARS-CoV-2 transmission on flights was very low ^23,24^. For example, no onward transmission was found on an airplane from China to Canada on January 22, 2020, carrying Canada’s first reported COVID-19 patient travelling from Wuhan ^25,26^. However, drawing robust conclusions from studies employing small samples is especially hard, as they are constrained by the availability of travel and epidemiological data by different transportation sources.

The potential spread of pathogens via transportation routes has also been reported and studied among other infectious diseases ^27,28^, such as severe acute respiratory syndrome (SARS) ^29-31^, tuberculosis ^32-35^, pandemic influenza A(H1N1) ^36-39^ and measles ^40^. However, results are not consistent across case-studies. Olsen et al. (2003) analysed the transmission risk of SARS on a flight carrying 120 passengers from Hong Kong to Beijing, and found that the attack rate reached 18.3%, and the risk to travellers sitting within 3 rows of the index patient was 3.1 times the risk than other locations ^31^. Nevertheless, Vogt et al. (2006) found that no SARS transmission occurred for a study involving 7 flights and 339 interviewees ^41^. Therefore, the overall transmission risk of infectious diseases on confined vehicles with high efficiency air filtration devices, e.g., airplane and high-speed train, remains unclear, and likely varies by disease.

Using anonymised individual itinerary and epidemiological data among confirmed COVID-19 patients and their close contacts on flights and high-speed trains across mainland China in the early days of the outbreak, we quantified the transmission risk of SARS-CoV-2 on airplanes and trains across seat locations and by joint travel (co-travel) time. To account for confounding relationships between passengers (e.g., family or friends) and their potential SARS-CoV-2 infections at home or working places before or after travel, we estimated the upper and lower bounds of transmission risk during the journey. The number of infected travellers was also estimated to understand the effectiveness of containment measures implemented across the country. The findings from this research can provide an improved evidence-base for tailoring travel interventions to reduce the transmission risk of COVID-19 in cabins and carriages during the resumption of socioeconomic activities and public transportation.

## Results

### COVID-19 prevalence among train and airplane passengers

A total of 9,265 airline passengers on 291 planes and 29,335 passengers on 830 G-trains (max speed 350 km/h) and D-trains (max speed 250 km/h) were included in this study during the early stage of COVID-19 outbreak in mainland China, from December 20, 2019 through March 17, 2020 (see data sources and enrolment criteria in Methods). Among them, 87.6% travelled via airplane and 96.9% travelled via train before the lockdown in the city of Wuhan on January 23, 2020 (Figure 1). Eventually, 336 airline travellers and 891 train passengers were later confirmed as COVID-19 cases within the 1-14 days following their travel. Further, 69.1% of cases travelling via airplane and 82.7% of cases travelling via train reported symptom onset before January 23, 2020 (e.g., during travel). However, only a small fraction of cases travelling via airplane (5.7%) and cases travelling via train (3.1%) had symptom onset after January 29, 2020, when all provinces of mainland China declared a first-level public health emergency response.

**Fig. 1.**
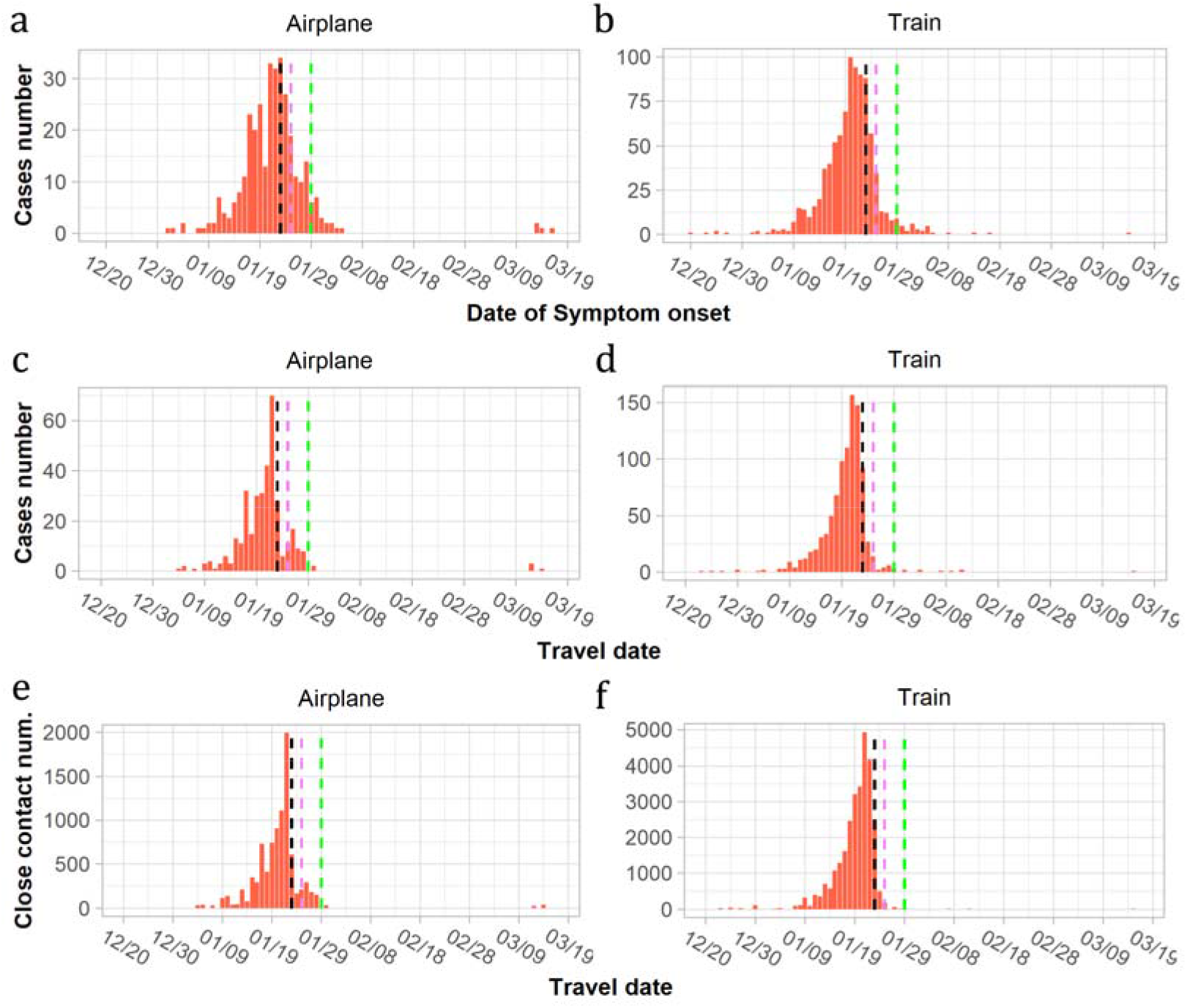
Number of passengers infected with COVID-19 and close contacts on airlines and trains across mainland China, December 2019 – March 2020. a-b. Number of cases by date of symptom onset; c-d. Number of cases by date of travel; e-f. Number of close contacts by date of travel. Black vertical dashed line: the date of Wuhan lockdown on January 23, 2020; Purple vertical dashed line: the Lunar New Year Day on January 25, 2020; Green vertical dashed line: the highest-level (Level 1) public health emergence response implemented across all provinces in mainland China, as of January 29, 2020.

### Attack rate of COVID-19 among passengers departing from Wuhan

175 index cases travelling by airplane were identified, with a further 587 index cases travelling via high-speed train from Wuhan before lockdown implementation (Table 1). To account for potential exposure to SARS-CoV-2 outside of the journey among family members or friends who were travelling together, we designed two assumptions for estimating the upper bound and lower bound of infection risks during travel, respectively (see Methods). For the former case, we assumed that all passengers were travelling independently during the journey, and without any previous contacts outside of the journey. In the latter assumption, we assumed that the index patient and passengers seating immediately adjacent to them with the same destinations were family or friends, and transmission of SARS-CoV-2 between them was therefore more likely to have occurred at home or in the workplace. Therefore, passengers seating immediately adjacent to an index patient were excluded from the attack rate (AR) calculation to estimate the potential lower bound of spreading risk of SARS-CoV-2 in airplanes and trains.

**Table 1.** COVID-19 transmission risk among airplane and high-speed train passengers departing from Wuhan.

| Characteristics | Airplane |  | High-speed train (G & D) |  |
| --- | --- | --- | --- | --- |
|  | Upper bound of risk | Lower bound of risk | Upper bound of risk | Lower bound of risk |
| Date of travel | 4 Jan 2020 ~ 23 Jan 2020 |  | 20 Dec 2019 ~ 23 Jan 2020 |  |
| Number of cases and close contacts |  |  |  |  |
| Total passengers | 5797 |  | 20276 |  |
| Close contacts | 5622 | 5400 | 19689 | 19300 |
| All cases | 209 | 209 | 656 | 656 |
| Index cases | 175 | 175 | 587 | 587 |
| Secondary cases | 34 | 18 | 69 | 41 |
| Number of airplanes/trains | 177 (Airbus: 67; Boeing: 110) |  | 506 (D: 214; G: 292) |  |
| Reproduction number (SD)<sup>a</sup> | 0.19 (0.45) | 0.10 (0.32) | 0.12 (0.40) | 0.07 (0.29) |
| Attack rate by seat location, 95% CI |  |  |  |  |
| Overall | 0.60% (34/5622),<br>0.43% - 0.84% | 0.33% (18/5400),<br>0.21% - 0.53% | 0.35% (69/19689),<br>0.28% - 0.44% | 0.21% (41/19300),<br>0.16% - 0.29% |
| Aisle seats | 0.56% (11/1974),<br>0.31% - 1.00% | 0.53% (10/1884),<br>0.29% - 0.97% | 0.34% (27/7894),<br>0.24% - 0.50% | 0.23% (18/7710),<br>0.15% - 0.37% |
| Middle seats | 0.71% (12/1692),<br>0.41% - 1.24% | 0.13% (2/1595),<br>0.03%-0.46% | 0.47% (16/3412),<br>0.29% - 0.76% | 0.21% (7/3320),<br>0.10% - 0.43% |
| Window seats | 0.56% (11/1956),<br>0.31% - 1.00% | 0.31% (6/1921),<br>0.14 - 0.68% | 0.31% (26/8383),<br>0.21% - 0.45% | 0.19% (16/8270),<br>0.12% - 0.31% |
| Seats adjacent to index cases | 9.20% (16/174),<br>0.57% - 14.4% | 1.32% (2/151),<br>0.37% - 4.70% | 3.63% (25/688),<br>2.47% - 5.31% | 0.88% (5/567),<br>0.38% - 2.04% |
| Seats not adjacent to index cases | 0.33% (18/5448),<br>0.21% - 0.52% | 0.30% (16/5249),<br>0.19% - 0.49% | 0.23% (44/19001),<br>0.17% - 0.31% | 0.19% (36/18733),<br>0.14% - 0.27% |

SD: standard deviation; 95%CI: 95% confidence interval. <sup>a</sup>The overall effective reproduction number during travel was estimated as the number of secondary cases divided by the number of index cases in each setting.

When estimating the upper bound of risk, 34 and 69 close contacts among passengers were considered to be infected on airplanes and trains departing from Wuhan, respectively, with an AR of 0.60% (34/5622, 95% confidence interval [CI] 0.43% - 0.84%) on planes and 0.35% (69/19689, 95%CI 0.28% - 0.44%) on trains (Table 1). The AR for high-speed trains was likely lower than that for airplanes (p = 0.01), with approximately 0.19 (SD 0.45) and 0.12 (SD 0.40) secondary cases per index case (effective reproduction number) during travel in cabins and carriages, respectively. The median time lag from departure to symptom onset was 4.0 days (interquartile range [IQR] 3.0 - 7.8) for airline passengers and 5.0 days (IQR 3.0 - 8.0) for train passengers. When estimating the lower bound of risk, the AR was 0.33% (18/5400, 95%CI 0.21% - 0.53%) on airplanes and 0.21% (41/19300, 95%CI 0.16% - 0.29%) on trains, and each index case infected 0.10 (SD 0.32) and 0.07 (SD 0.29) passengers on airplanes and high-speed trains, respectively. Additionally, the AR on high-speed trains was likely lower than that on airplane departing from Wuhan or other cities in mainland China (Extended Data Table 1) (p < 0.01).

### COVID-19 transmission risk by seat locations

The AR varied greatly by seat within three rows and six columns to the index patient seat (Figure 2). In the upper-bound risk estimation, the seats immediately adjacent to the index cases had the highest risk, with an AR of 9.2% (95%CI 5.7% - 14.4%) on airplanes and 3.6% (95%CI 2.5% - 5.3%) on trains. Compared to other seats, the relative risk (RR) of these seats was 27.8 (95%CI 14.4 - 53.7) on airplanes and 15.7 (95%CI 9.7 - 25.5) on trains. Generally, passengers seated on the same row as the index case had a higher risk than passengers at other rows, with a RR of 10.6 (95%CI 5.3 - 21.1) for airplanes and 8.9 (95%CI 5.5 - 14.3) for trains, respectively. However, ARs for the seats at one to three rows apart from the index patient showed no significant difference between each other (p > 0.05).

**Fig. 2.**
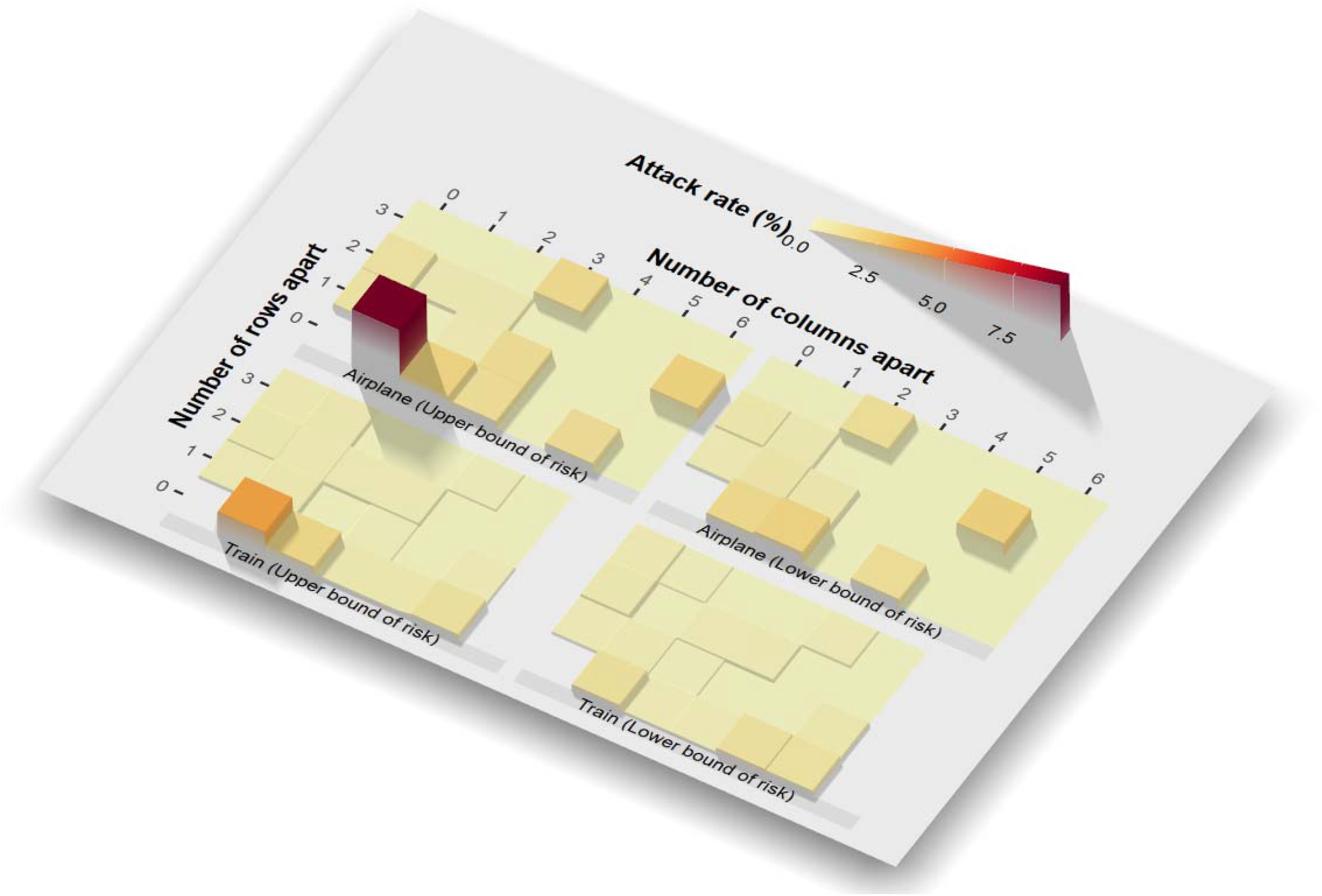
Attack rates of COVID-19 per different seats apart from index cases on airplanes and high-speed trains for passengers departing from Wuhan, as of January 23, 2020. The seats of index cases were indicated at (row 0, column 0).

**Fig. 3.**
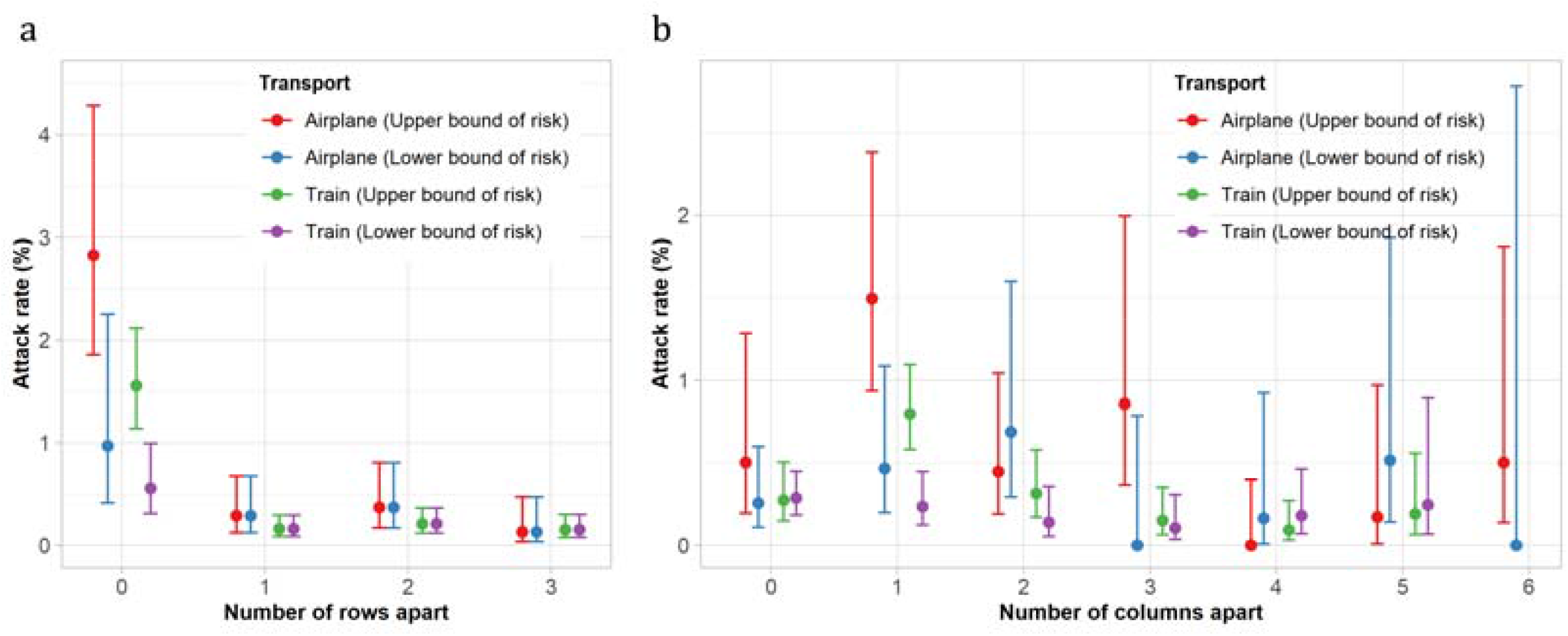
Attack rates of COVID-19 on airplane and high-speed trains for passengers departing from Wuhan, as of January 23, 2020. a. Attack rate of seats by row apart from the index patient; b. Attack rate of seats by column apart from the index patient.

**Fig. 4.**
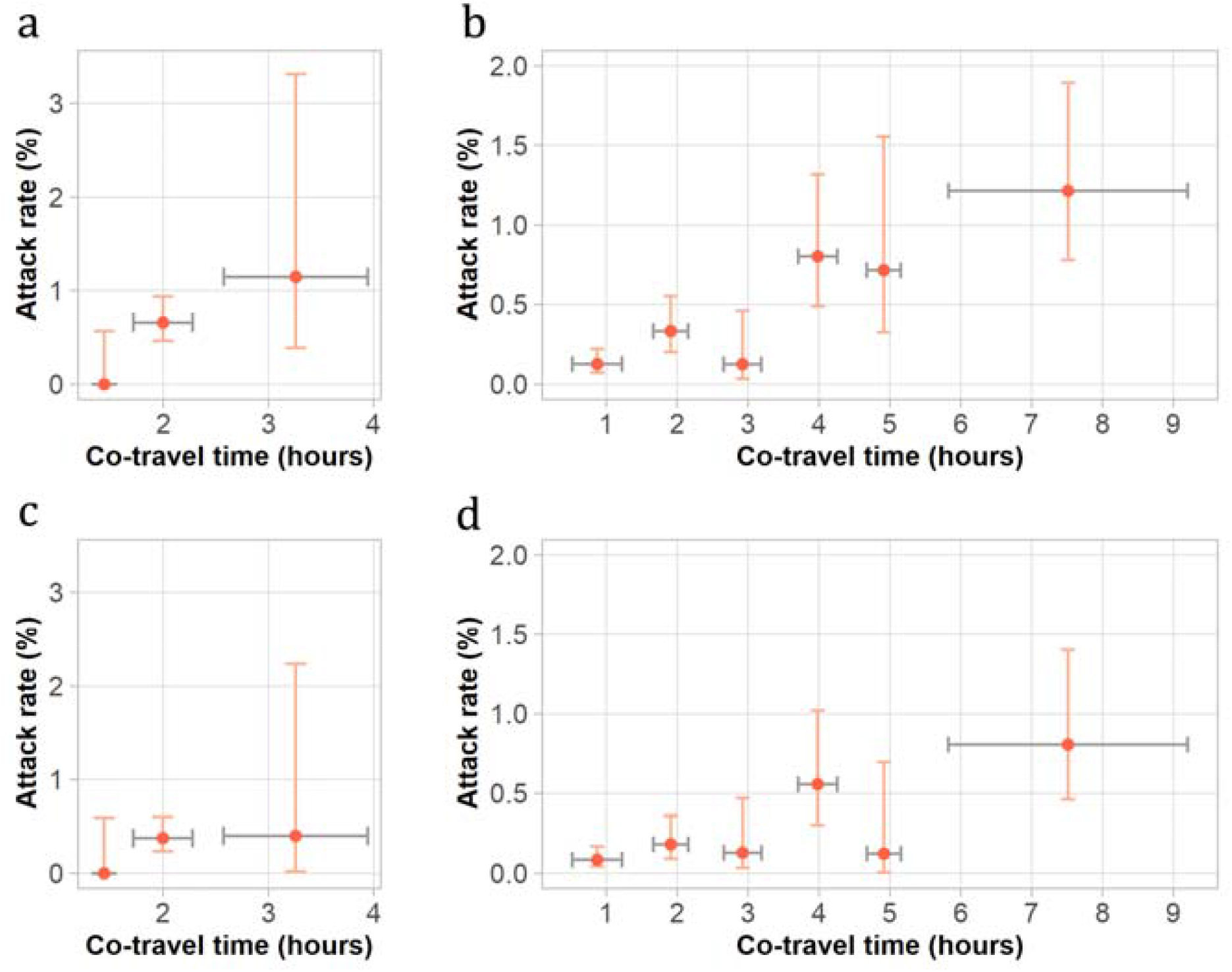
Relationships between COVID-19 attack rate and co-travel time on airplane and trains departing from Wuhan, as of January 23, 2020. a-b. Upper bounds of attack rates on airplanes (a) and high-speed trains (b), respectively; c-d. Lower bounds of attack rates on airplanes (c) and high-speed trains (d), respectively. Vertical bars show 95% confidence interval of attack rate estimates; horizontal bars represent the standard deviation of co-travel time.

In terms of seat location on airplanes, i.e., window, middle and aisle seats, the middle seat had the highest AR (0.7%, 95%CI 0.4% - 1.2%), followed by the window seat (0.6%, 95%CI 0.3% - 1.0%) and the aisle seat (0.6%, 95%CI 0.3% - 1.0%) for the upper bound estimation, likely due to the fact that middle seats are exposed to individuals on either side, where window and aisle seats are exposed to only one individual. For trains, the middle seat also had the highest AR 0.4% (95%CI 0.3% - 0.8%). No significant difference in AR was found on seats between Airbus and Boeing airplanes (p = 1.0), and between G-trains and D- trains (p = 0.64) (Extended Data Table 2).

### Effect of co-travel duration on COVID-19 transmission risk

Travel time among airplane passengers ranged from 1.1 to 4.3 hours (mean 2.0, SD 0.5), with approximately 98% travelling less than 3 hours. On average, the upper bound of AR increased from 0.7% (95%CI 0.5% - 1.0%) to 1.2% (95%CI 0.4% - 3.3%) when the co-travel time increased from 2.0 hours to 3.3 hours (Extended Data Table 3), while the lower bound estimates of AR increased from a relatively low risk (0.0%, 95%CI 0.0% - 0.6%) within 1.5 hours, to 0.4% (95%CI 0.02% - 2.2%) for a joint travel duration of 3.3 hours. However, due to the small number of secondary cases in different groups of co-travel time intervals, there was no significant difference between the estimated upper and lower bounds of ARs (p = 0.06).

The travel time of train passengers ranged from 0.2 to 12.6 hours, with a mean of 2.2 hours (SD 2.0), and 91% passengers travelling less than 5 hours. Within 3.5 hours’ co-travel, the average ARs had an upper bound of 0.19% (95%CI 0.1% - 0.3%) and lower bound of 0.1% (95%CI 0.07% - 0.2%). When the co-travel duration exceeded 3.5 hours, the ARs increased to 0.9% (95%CI 0.7% - 1.3%) and 0.6% (95%CI 0.4% - 0.8%), respectively, which were significantly higher than the risks within 3.5 hours’ co-travel (p < 0.01), with RRs of 5.0 (95%CI 3.1 - 8.0) and 4.8 (95%CI 2.6 - 8.8), respectively. The *q* index for measuring spatial heterogeneity of risk (see Methods) was 0.33 (p = 0.02), taking the row number and hourly co-travel time as the unit of stratification.

### Impact of non-pharmaceutical interventions

Non-pharmaceutical interventions were widely implemented across China following Wuhan’s lockdown on January 23, 2020, including measures such as case detection and isolation, travel restrictions, temperature screening at airports and train stations, mandatory face coverings, personal hygiene protection, and more ^42,43^. As a result, case numbers and passengers travelling by both airplane and high-speed rail decreased drastically throughout mainland China, particularly after the implementation of intensive public health emergence response efforts (i.e., the highest level interventions and travel restrictions) across China on January 29, 2020 (Figure 1; Extended Data Table 4 and 5).

## Discussion

Although many studies have explored the impact of human mobility and relevant interventions on the spread of infectious diseases ^2,3,44^, our study is the first to quantify the risk of SARS-CoV-2 transmission among individual passengers on both airplanes and trains across China. These findings inform transmission probability estimates of COVID-19 in confined environments, which may help to formulate targeted strategies to protect travellers and control long-distance transmission of the disease. Based on individual case and passenger data on airplanes and high-speed trains, our research showed that overall transmission risk of SARS-CoV-2 among passengers in the early days of outbreaks was relatively low via air and rail travel. We further quantified the relationship between transmission risk and other travel factors, such as seat distance from an infectious individual and co-travel time.

We found that transmission of SARS-CoV-2 in cabins and carriages might have occurred, but the overall risk to a given individual was relatively low for domestic air and train travel, even under upper bound risk assumptions. Several factors might explain this; firstly, individuals are less likely to travel after illness onset, in particular severe infections. Secondly, the degree of infectivity among travellers may vary temporally. For example, some travellers infected with SARS-CoV-2 might be pre-symptomatic and still in the latent period of infection, defined as the period between exposure to the virus and the onset of infectiousness. Thirdly, modern commercial airplanes and high-speed trains are commonly equipped with high efficiency particulate air filters (HEPA), which can effectively filter out 99.9% of dust, bacteria and viruses with a particle size of more than 0.3μm ^40^, thereby reducing transmission risk of respiratory infectious diseases ^45-48^. When the air enters the passenger cabin, it undergoes high temperature disinfection and cooling. Aircraft cabin air conditioners can complete up to 20 air changes per hour, reducing the presence of virus-containing droplets and the risk of disease transmission in the cabin. Towards this, we found that the difference in risk was not significant by aircraft and train type within our analyses.

Our findings suggest risk varied with joint travel time and seat distance to the index case, with risk increasing by longer travel time. The effect of co-travel duration to COVID-19 transmission risk was more obvious among train passengers than domestic air travel, as there were more passengers travelling for longer periods of time by high-speed trains across the country. Additionally, there are several factors which may explain the higher risk we observed among travellers seated in the same row as index patients. Firstly, droplets and aerosols produced by the index patient’s talking, coughing, and sneezing might be more likely to spread within the immediate vicinity, such as the same row ^49,50^. Further, the seat backs between the front and rear rows might partially block the spread of the virus in the forward and backward positions. Secondly, each row of seats has an independent air-conditioning exhaust system that blows air from top to bottom and is filtered away from the bottom of the seat. This design could reduce the speed of air diffusion to the surroundings and thereby dilute the virus concentration ^32^. Thirdly, passengers in the same row might have more opportunities for social contact, especially if they are travelling with family members or friends. Lastly, non-pharmaceutical interventions that were in place across China since late January 2020 have effectively contained the spread and further transmission of COVID-19 across the country ^42,43^, subsequently reducing the infection risk among travellers. Further containment measures such as temperature screening at stations or airports, face mask usage, and travel restrictions for high risk areas might work synergistically to minimize transmission risk in transit ^51^.

Despite our findings, other studies have reported a high transmission risk of other infectious diseases on planes ^27,28^. Kenyon et al. (1996) studied the transmission risk of tuberculosis on an aircraft that flew for 8.5 hours and found that the attack rate within and outside of two seat rows to the index patient was 30.8% and 3.6%, respectively ^34^. Hertzberg and Weiss (2016) analysed the average risk of SARS, H1N1, measles and other infectious diseases on the airplane, and concluded that infection risk within and outside of two rows of the index patient was 6% and 2%, respectively ^52^. However, according to the analysis of data from hundreds of routes in this study, the average AR of COVID-19 within three rows of the index patient was much lower than that for other diseases reported in previous results. These findings suggest that the transmission risks and mechanisms of COVID-19 and other diseases on airplanes and trains are complicated, accompanied by certain random factors and super spreader events ^19,31,53^.

Our study was subject to a variety of limitations. Firstly, due to the limitation of data availability, we only studied the risk of transmission within three rows (7 rows in total) in front of and behind the index case, and therefore did not consider possible transmission within a larger scope (including flight and train crews). However, previous studies showed that the seats within 3 rows to the index patients had the highest risk ^52^. Secondly, as with other similar studies, we cannot accurately determine when and where the infection occurred due to recall bias from passenger reported information^32^. Cases may have been infected in other places before or after travel, such as in terminals or railway stations, during the boarding process or even outside the airport and station. This might overestimate the risk of spreading COVID-19 on planes or trains, although we accounted for this by employing two scenarios to estimate the interval of risk. Thirdly, data on usage of personal protective equipment such as masks and goggles during the flight were not available to account for the impact of these interventions, although few passengers might use these measures before January 23, 2020 when the lockdown of Wuhan was in place. Fourthly, asymptomatic travellers might not be captured in this study, and transmission risk before the symptom onset might be overlooked. Lastly, the difference in the size and cabin layout of aircrafts and trains was not considered in the study, where first class cabins tend to have more space between passengers than economy cabins. This information should be taken into account in future research. However, our findings can improve our understanding of SARS-CoV-2 transmission during travel and support countries in tailoring interventions to minimize the risk in cabins and carriages during the resumption of socioeconomic activities and public transportation.

## Data Availability

The data and code for plotting figures in this study have been made openly available for further use at https://github.com/maoguihu/TransmisionRiskOnAirplane-Train. The individual itinerary data of cases and passengers included in the current study are not publicly available since this would compromise the agreement with the data provider, but the information on the process of requesting access to the data that support the findings of this study are available from the correspondence author.

## Methods

### Data sources

Since the COVID-19 outbreak has been first reported, the national and local Centres for Disease Control and Prevention (CDC) across China have conducted intensive epidemiological investigations to identify COVID-19 cases and close contacts as well as their travel history. We included all confirmed COVID-19 cases who had a travel history of domestic flight or high-speed train during illness or within 14 days before symptom onset, as of March 17, 2020. A total of 291 airplanes and 830 high-speed trains with confirmed COVID-19 cases on board were enrolled in this study. The date of travel, flights/trains no., seat no., departure and arrival airports or stations of cases and passengers as potential close contacts were collected to analyse the potential infectious risk of the disease. To measure the duration of travel, we searched flight timetables and flight types at the Variflight (www.variflight.com) and the Ctrip (www.ctrip.com). Railway timetables were obtained from the China Railway’s booking website (www.12306.cn). The flight dataset contained a total of 291 flights flying between 81 cities from January 4, 2020 to March 14, 2020, of which Airbus and Boeing accounted for 43.6% (127) and 55.3% (161) respectively, with 234 flights departing from Wuhan before the lockdown on January 23, 2020. A description of the dataset of high-speed G (max speed 350 km/h) trains has also been used and detailed in our previous preliminary study ^22^, but in this research, we included both G and D (max speed 250 km/h) high-speed trains that have similar seat arrangement and air-conditioning filtration system ^54^. The high-speed train dataset contained 830 trains, including 328 (39.5%) D trains and 502 (60.5%) G trains, stopped at 582 stations from December 20, 2019 up until March 17, 2020. Additionally, there were a total of 506 trains travelled from or stopped by stations in Wuhan before its lockdown.

### Definitions of case and close contact

According to the Diagnosis and Treatment Scheme for COVID-19 released by the National Health Commission of China ^55^, a confirmed case was diagnosed with a positive result for SARS-CoV-2 nucleic acid by real-time reverse transcriptase–polymerase chain reaction (RT-PCR) assay or high-throughput sequencing of nasal and pharyngeal swab specimens. On an airplane or each coach of a train, a passenger was defined as an index case if he/she: i) was confirmed with COVID-19 after the travel; and ii) had symptom onset within 2 days before travel or within 14 days after travel; and iii) had the earliest date of symptom onset among passengers within 3 rows in the case where there was more than one case. Passengers within 3 rows of the index case seat were considered to be close contacts for estimating the upper bound of risk detailed below. Given an average incubation period of 5 days (range 2-14 days), secondary COVID-19 cases were defined as cases among close contacts who had symptom onset within 2 to 14 days later than the index case after travel ^56^.

The relationship and contacts between passengers were important factors in determining the transmission risk of COVID-19 on airplanes and trains, because family members, colleagues or friends frequently co-travel and might have a higher transmission risk at home or in the workplace, as opposed to travel ^57^. However, individual social network information was not available for this study. Therefore, to account for group travel among family or friends, we used two assumptions for estimating the upper bound and lower bound of infection risks on transport, respectively. For estimating the upper-bound risk, we assumed that there was no family or friend relationship between travellers, nor any contacts before and after the journey, to estimate the potential high risks (upper bound) of SARS-CoV-2 transmission during air and train travel. For estimating the lower bound of risk, passengers were assumed to be travelling with their family members or friends if a small group of passengers included one index COVID-19 patient, and passengers seated immediately adjacent to this index patient shared the same departure and destination. Therefore, passengers fitting this definition were excluded from close contacts of the index patient to estimate the potential low risks (lower bound) of SARS-CoV-2 spread on airplanes and trains, respectively.

### Assessing transmission risks

The attack rate (AR) of a seat was calculated as the number of confirmed cases divided by the total number of passengers that used the same seat. Relative risk (RR) and Chi-squared tests were used to compare the risks between different seats. Fisher’s exact test was used when the Chi-squared approximation might be incorrect because of small sample sizes. The 95% confidence interval (CI) of estimated AR was estimated using Wilson’s binomial method. Risk ratio was calculated using Wald’s unconditional maximum likelihood estimation, and its 95% CI was estimated using Wald’s normal approximation. The number of secondary cases infected per index case during travel was also used as an effective reproduction number of COVID-19 transmission among passengers on airplanes and trains, respectively. Additionally, to understand the risk of SARS-CoV-2 transmission among travellers in transit before the wide implementation of non-pharmaceutical interventions in the early days of the outbreak, we focused on the individual-level transmission risk of COVID-19 among passengers on domestic flights and high-speed trains, departing from Wuhan before the lockdown was implemented on January 23, 2020.

### Measuring spatial heterogeneity of risk

The seat distance between each close contact and the index case is an important factor in disease transmission. Wang’s *q* index was adopted to measure the spatial heterogeneity in ARs between rows and columns of seats ^58,59^. The *q* index varies between 0 and 1. The closer the *q* value is to 1, the stronger the heterogeneity of the spatial distribution of transmission risk.

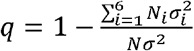

where *N* and *σ*^2^ stand for the number of seats and the variance of attack rates for all considered seats, respectively. *N*_*i*_ and 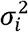 denote the number of seats and the variance of attack rates for seats at the *i*^th^ row, respectively.

### Assessing the impact of co-travel time on COVID-19 transmission

The duration of domestic flights departing from Wuhan was mostly around two to three hours, and generally within four hours. To measure the relationship between co-travel time with the COVID-19 transmission risk on board, we grouped the co-travel times of airline passengers with an index case into three categories: 0 – 1.5 hours, 1.5 – 2.5 hours, and more than 2.5 hours. Similarly, the duration of co-travel on high-speed train was classified into six groups: 0 – 1.5 hours, 1.5 – 2.5 hours, 2.5 – 3.5 hours, 3.5 – 4.5 hours, 4.5 – 5.5 hours, and more than 5.5 hours. Chi-squared tests were used to compare the risk between different co-travel time groups. All statistical analyses in this study were conducted using R version 4.0.3 (R Foundation for Statistical Computing, Vienna, Austria).

## Acknowledgments

We thank staff members at disease control institutions, hospitals, and health administrations across China for field investigation, administration, and data collection. We also thank Prof Marc Lipsitch at the Harvard T.H. Chan School of Public Health for his insightful comments to improve this paper. This study was supported by the National Science and Technology Major Project of China (2017ZX10201302 and 2016ZX10004222-009), the National Natural Science Foundation of China (41771434, 41531179, and 81773498), and the Bill & Melinda Gates Foundation (OPP1134076 and INV-024911).

## Author contributions

MH designed the study, built the model, collected data, finalised the analysis, interpreted the findings, and wrote the manuscript. SL conceived the study, built the model, interpreted the findings, wrote and revised the manuscript. JW conceived and designed the study, interpreted the findings, and commented on and revised drafts of the manuscript. HL, CX, BM, XZ, YF, and QY collected data, interpreted the findings, and revised drafts of the manuscript. CWR, AC, JRF, NWR, ZL, WY, and AJT interpreted the findings, and commented on and revised drafts of the manuscript. All authors read and approved the final manuscript.

## Ethical approval

The collection and analysis of COVID-19 case data were determined by the National Health Commission of China to investigate and control the outbreak. Ethical clearance for collecting and using secondary data in this study was granted by the institutional review board of the University of Southampton (No. 61865). All data were supplied and analysed in an anonymous format, without access to personal identifying information.

## Data and code availability

**Extended Data Table 1.**
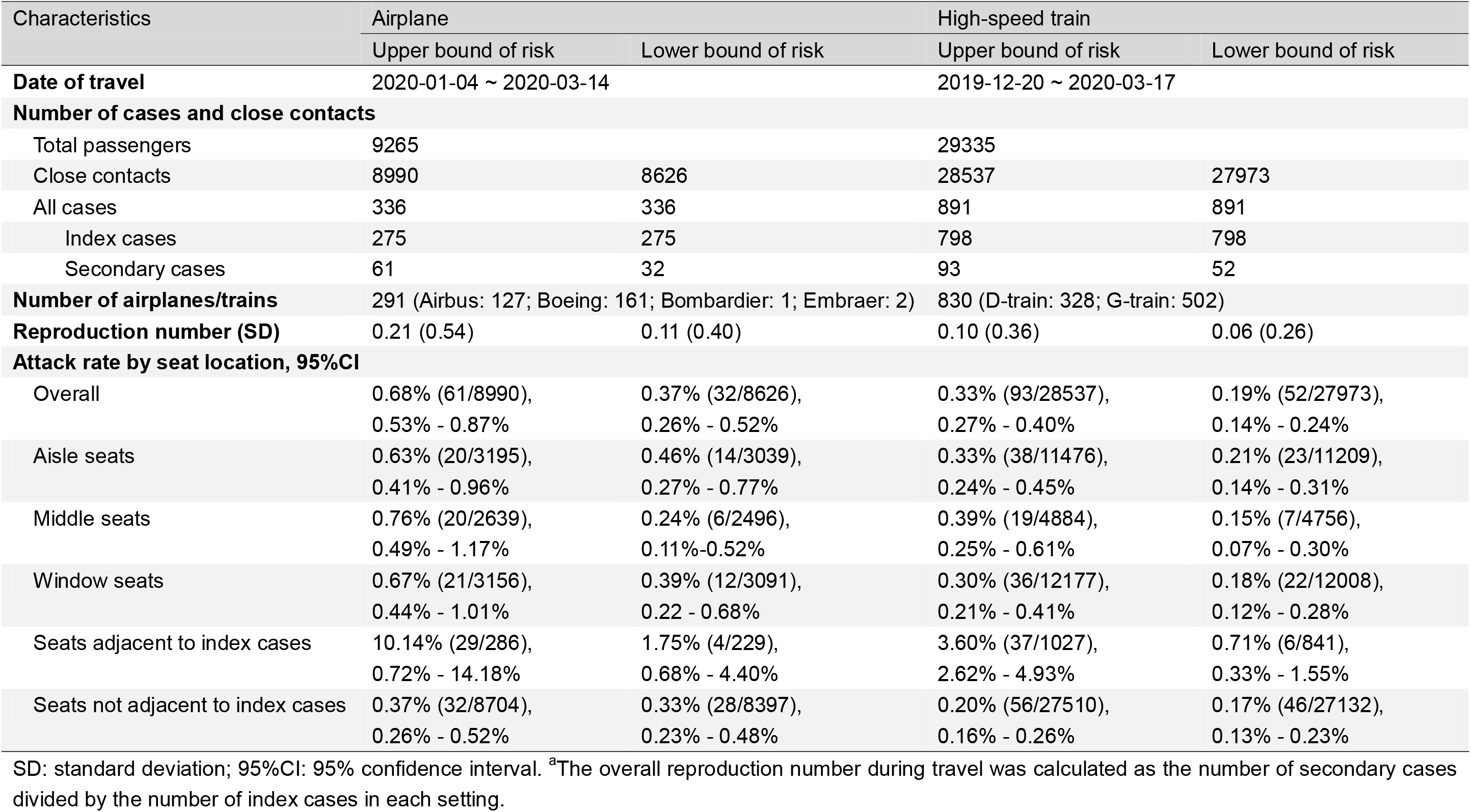
COVID-19 transmission risk among airplane and high-speed train passengers in mainland China.

| Characteristics | Airplane |  | High-speed train |  |
| --- | --- | --- | --- | --- |
|  | Upper bound of risk | Lower bound of risk | Upper bound of risk | Lower bound of risk |
| Date of travel | 2020-01-04 ~ 2020-03-14 |  | 2019-12-20 ~ 2020-03-17 |  |
| Number of cases and close contacts |  |  |  |  |
| Total passengers | 9265 |  | 29335 |  |
| Close contacts | 8990 | 8626 | 28537 | 27973 |
| All cases | 336 | 336 | 891 | 891 |
| Index cases | 275 | 275 | 798 | 798 |
| Secondary cases | 61 | 32 | 93 | 52 |
| Number of airplanes/trains | 291 (Airbus: 127; Boeing: 161; Bombardier: 1; Embraer: 2) |  | 830 (D-train: 328; G-train: 502) |  |
| Reproduction number (SD) | 0.21 (0.54) | 0.11 (0.40) | 0.10 (0.36) | 0.06 (0.26) |
| Attack rate by seat location, 95%CI |  |  |  |  |
| Overall | 0.68% (61/8990), | 0.37% (32/8626), | 0.33% (93/28537), | 0.19% (52/27973), |
|  | 0.53% - 0.87% | 0.26% - 0.52% | 0.27% - 0.40% | 0.14% - 0.24% |
| Aisle seats | 0.63% (20/3195), | 0.46% (14/3039), | 0.33% (38/11476), | 0.21% (23/11209), |
|  | 0.41% - 0.96% | 0.27% - 0.77% | 0.24% - 0.45% | 0.14% - 0.31% |
| Middle seats | 0.76% (20/2639), | 0.24% (6/2496), | 0.39% (19/4884), | 0.15% (7/4756), |
|  | 0.49% - 1.17% | 0.11%-0.52% | 0.25% - 0.61% | 0.07% - 0.30% |
| Window seats | 0.67% (21/3156), | 0.39% (12/3091), | 0.30% (36/12177), | 0.18% (22/12008), |
|  | 0.44% - 1.01% | 0.22 - 0.68% | 0.21% - 0.41% | 0.12% - 0.28% |
| Seats adjacent to index cases | 10.14% (29/286), | 1.75% (4/229), | 3.60% (37/1027), | 0.71% (6/841), |
|  | 0.72% - 14.18% | 0.68% - 4.40% | 2.62% - 4.93% | 0.33% - 1.55% |
| Seats not adjacent to index cases | 0.37% (32/8704), | 0.33% (28/8397), | 0.20% (56/27510), | 0.17% (46/27132), |
|  | 0.26% - 0.52% | 0.23% - 0.48% | 0.16% - 0.26% | 0.13% - 0.23% |

SD: standard deviation; 95%CI: 95% confidence interval. <sup>a</sup>The overall reproduction number during travel was calculated as the number of secondary cases divided by the number of index cases in each setting.

**Extended Data Table 2.** **Attack rate by the type of airplanes and high-speed trains**.

| Type |  | Upper bound of risk | Lower bound of risk |
| --- | --- | --- | --- |
| Airplane | Airbus | 0.61% (13/2119) | 0.34% (7/2032) |
|  |  | 0.36% - 1.05% <sup>a</sup> | 0.17% - 0.71% <sup>a</sup> |
|  | Boeing | 0.60% (21/3503) | 0.33% (11/3368) |
|  |  | 0.39% - 0.91% <sup>a</sup> | 0.18% - 0.58% <sup>a</sup> |
| High-speed train | D-train | 0.33% (27/8165) | 0.19% (15/7995) |
|  |  | 0.23% - 0.48% <sup>a</sup> | 0.11% - 0.31% <sup>a</sup> |
|  | G-train | 0.36% (42/11524) | 0.23% (26/11305) |
|  |  | 0.27% - 0.49% <sup>a</sup> | 0.16% - 0.34% <sup>a</sup> |

<sup>a</sup> 95% confidence interval.

**Extended Data Table 3.** Attack rate (%) by co-travel time among airplane and high-speed train passengers departing from Wuhan.

| Type | Co-travel time, hours (mean) | Upper bound of risk (95%CI) | Lower bound of risk (95%CI) |
| --- | --- | --- | --- |
| Airplane | 1.1-1.5 (1.4) | 0 (0/669, 0-0.57) | 0 (0/641, 0-0.60) |
|  | 1.5-2.5 (2.0) | 0.66 (31/4691, 0.47-0.94) | 0.38 (17/4510, 0.20-0.60) |
|  | 2.5-4.3 (3.3) | 1.15 (3/262, 0.39-3.31) | 0.40 (1/249, 0.02-2.24) |
| Train | 0.2-1.5 (0.9) | 0.13 (12/9375, 0.07-0.22) | 0.09 (8/9290, 0.04-0.17) |
|  | 1.5-2.5 (1.9) | 0.34 (15/4470, 0.20-0.55) | 0.18 (8/4374, 0.09-0.36) |
|  | 2.5-3.5 (2.9) | 0.13 (2/1578, 0.03-0.46) | 0.13 (2/1545, 0.04-0.47) |
|  | 3.5-4.5 (4.0) | 0.80 (15/1867, 0.49-1.32) | 0.56 (10/1795, 0.30-1.02) |
|  | 4.5-5.5 (4.9) | 0.72 (6/838, 0.33-1.55) | 0.12 (1/809, 0.01-0.70) |
|  | 5.5-12.6 (7.5) | 1.22 (19/1561, 0.78-1.89) | 0.81 (12/1487, 0.46-1.41) |

**Extended Data Table 4.** COVID-19 transmission risk among airplane and high-speed train passengers in mainland China from January 24 to January 28, 2020.

| Characteristics | Airplane |  | High-speed train |  |
| --- | --- | --- | --- | --- |
|  | Upper bound of risk | Lower bound of risk | Upper bound of risk | Lower bound of risk |
| **Number of cases and close contacts** |  |  |  |  |
| Total passengers | 1051 |  | 816 |  |
| Close contacts | 1010 | 963 | 776 | 747 |
| All cases | 51 | 51 | 53 | 53 |
| Index cases | 41 | 41 | 40 | 39 |
| Secondary cases | 10 | 6 | 13 | 4 |
| **Number of airplanes/trains** | 41 (Airbus: 23; Boeing: 16; Embraer: 2) |  | 41 (D-train: 16; G-train: 25) |  |
| **Overall attack rate, 95%CI** | 0.99% (10/1010),<br>0.54% - 1.81% | 0.66% (6/963),<br>0.29% - 1.35% | 1.67% (13/776),<br>0.98% - 2.85% | 0.54% (4/747),<br>0.21% - 1.37% |

SD: standard deviation; 95%CI: 95% confidence interval.

**Extended Data Table 5.** COVID-19 transmission risk among airplane and high-speed train passengers in mainland China from 29 January 2020 up until 17 March 2020.

| Characteristics | Airplane |  | High-speed train |  |
| --- | --- | --- | --- | --- |
|  | Upper bound of risk | Lower bound of risk | Upper bound of risk | Lower bound of risk |
| **Number of cases and close contacts** |  |  |  |  |
| Total passengers | 101 |  | 103 |  |
| Close contacts | 96 | 92 | 91 | 88 |
| All cases | 6 | 6 | 13 | 13 |
| Index cases | 5 | 5 | 12 | 11 |
| Secondary cases | 1 | 1 | 1 | 1 |
| **Number of airplanes/trains** | 4 (Airbus: 2; Boeing: 2) |  | 13 (D-train: 5; G-train: 8) |  |
| **Overall attack rate, 95%CI** | 1.00% (1/96),<br>0.18% - 5.67% | 1.09% (1/92),<br>0.19% - 5.90% | 1.10% (1/91),<br>0.19% - 5.96% | 1.13% (1/88),<br>0.20% - 6.16% |

SD: standard deviation; 95%CI: 95% confidence interval. The highest-level intensive public health emergence responses had been implemented across all provinces in mainland China since 29 January 2020.

**Extended Data Fig. 1.**
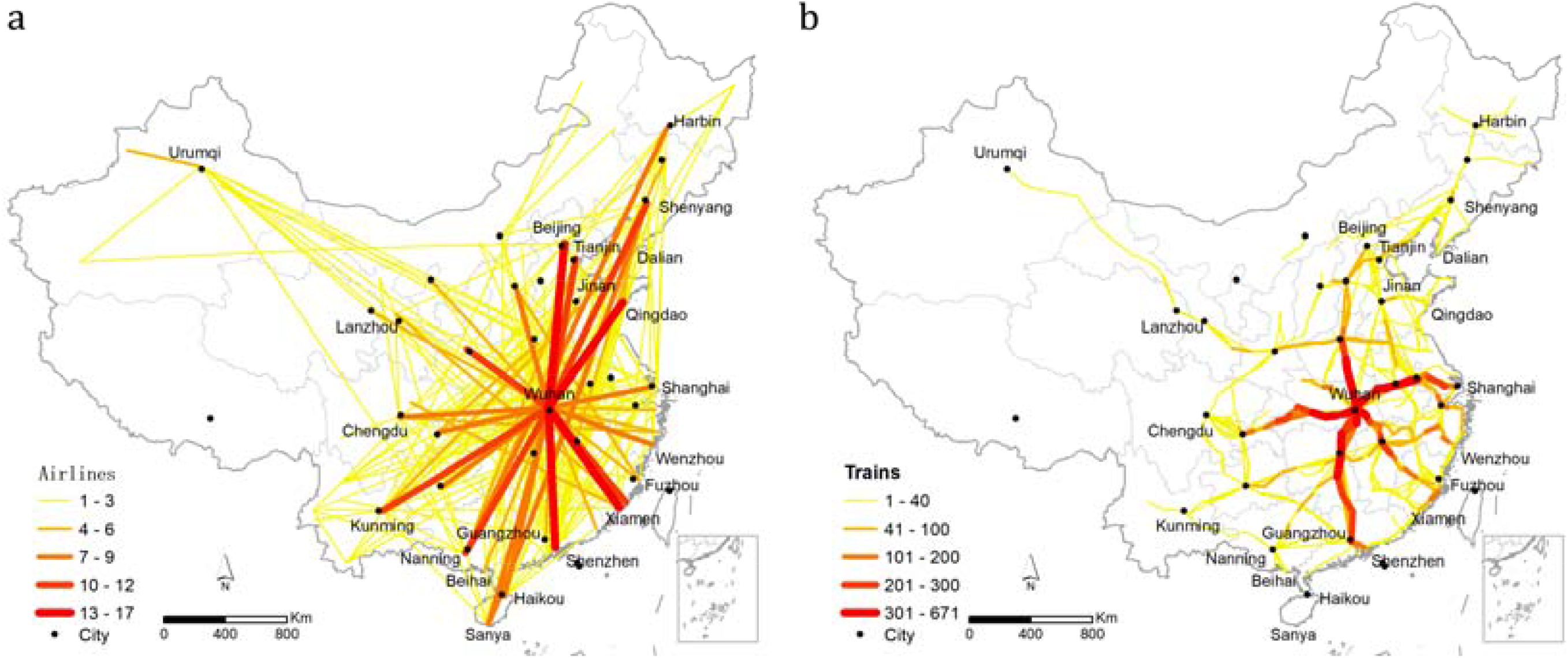
Routes and numbers of domestic airlines (a) and high-speed trains (b) from Wuhan before its lockdown on January 23, 2020.

**Extended Data Fig. 2.**
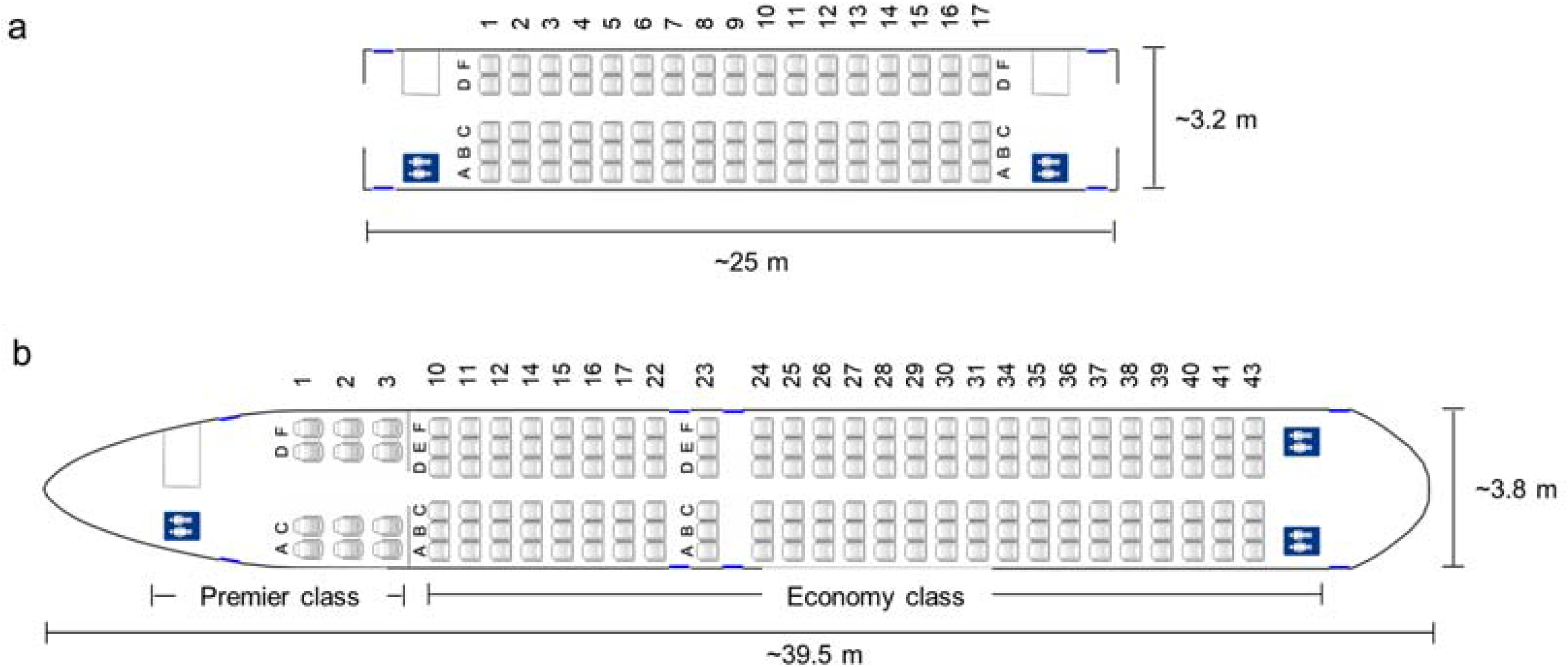
Distribution of seats in a train and airplane (A) Distribution of second-class seats in a typical high-speed train coach. (B) Distribution of seats in a typical airplane. The columns are represented by letters, and the rows are indicated by numbers.

**Extended Data Fig. 3.**
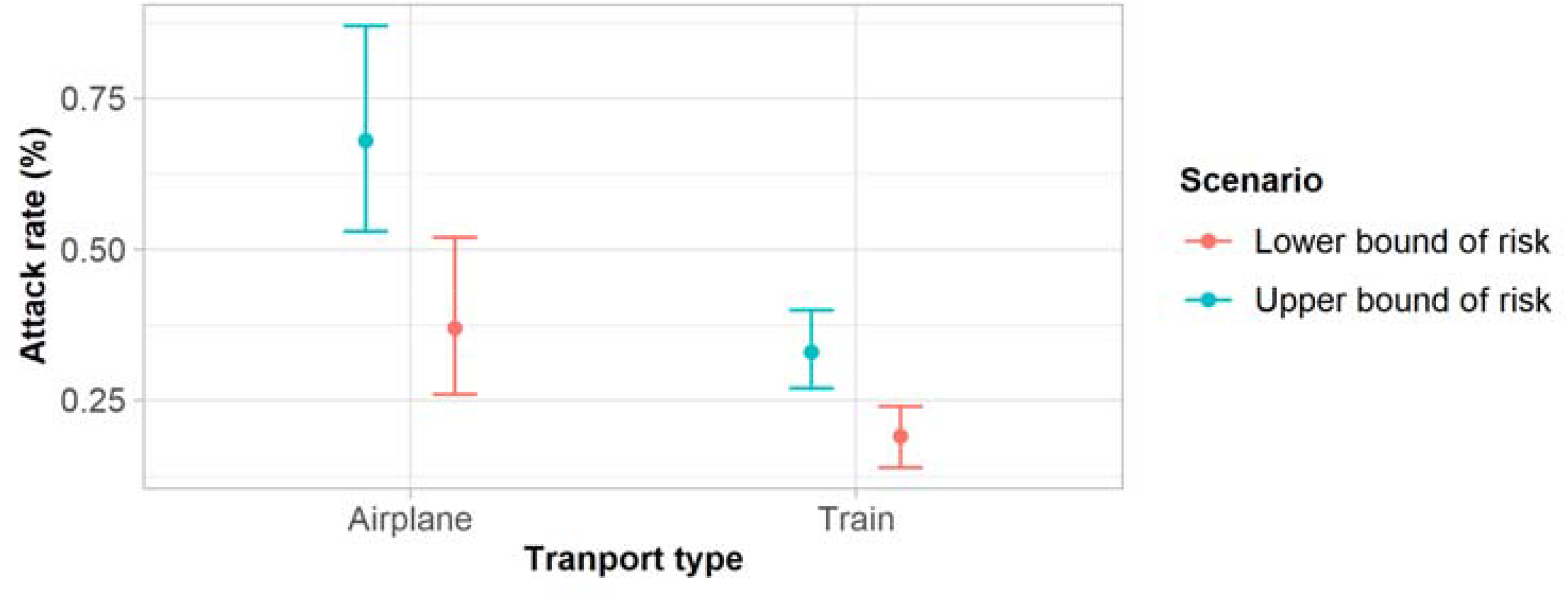
Overall AR on airplane and high-speed train in mainland China. (Vertical bars denote 95% confidence interval of attack rate.)

